# Prevalence and impact of SARS-CoV-2, influenza, respiratory syncytial virus (RSV) infection and respiratory illness on UK healthcare workers during winter 2023/24 (September 2023 to March 2024): SIREN cohort study

**DOI:** 10.1101/2025.03.07.25323556

**Authors:** Sarah Foulkes, Katie Munro, Dominic Sparkes, Jameel Khawam, Sophie Russell, Angela Dunne, Jean Timeyin, Nipunadi Hettiarachchi, Michelle Cairns, Declan T Bradley, Elen De Lacy, Kevin Wilson, Nick Andrews, Andre Charlett, Respiratory Virus and Microbiome Initiative (RVI) team, Katie Bellis, Ya-Lin Huang, Matthew Forbes, Andrea Frick-Kretschmer, Marissa Knoll, Ewan M. Harrison, SIREN study group, Colin S Brown, Ana Atti, Jasmin Islam, Susan Hopkins, Victoria Hall

## Abstract

During the winter, healthcare systems experience additional pressures due to increases in respiratory infections and staff absence. We aimed to determine the prevalence of respiratory viruses and impact on sickness absence in the SIREN healthcare worker (HCW) cohort during winter 2023/24.

SIREN is a cohort study with linked testing, vaccination, demographic, symptoms and sick leave data. Participants undergo fortnightly multiplex PCR testing for SARS-CoV-2, influenza and RSV, regardless of symptoms. The proportion of participants who took sick leave, the total number and median of sick leave days taken was calculated by viral infection and vaccination status. Logistic regression was used to estimate the association between sick leave and vaccination status.

5,287 participants were included, 78·3% female, median age 54 years. There were 1,828 infections among 1,659 participants (1,161 SARS-CoV-2; 387 RSV; 280 influenza infections). Influenza and RSV peaked in December (1·2%; 1·9%), SARS-CoV-2 peaked in September and December 2023 (4·0%; 4·3%).

Regardless of a known infection, 35.8% (1,892/5,287) took sick leave, resulting in 10,168 days (median 5 days per person; range 1-98 days). Vaccination was not significantly associated with reduced sick leave (adjusted odd ratios 0·98; 95%CI 0·87, 1·10).

Respiratory illness caused a substantial burden on the SIREN healthcare workforce over winter, with all three viruses contributing. Reduced number of staff at work and pressures to work through illness have implications for healthcare resilience.

## INTRODUCTION

Increased circulation of respiratory viruses each winter causes significant strain on healthcare systems, due to upsurges in both numbers of patients and in staff sickness absence (1, 2), commonly referred to as “Winter Pressures”. Pre-COVID-19 pandemic, influenza was the leading contributor to cases of acute respiratory illness however since the pandemic, SARS-CoV-2 has become a major contributor over winter (3-5). All three viruses (influenza, SARS-CoV-2 and Respiratory Syncytial Virus (RSV)) can cause a range of clinical presentations and contribute to significant morbidity and mortality in the population, with severe outcomes most common at extremes of age and in immunocompromised individuals (6-8).

Seasonal influenza and COVID-19 vaccination campaigns amongst healthcare workers (HCWs) aim to reduce staff sickness absence and nosocomial transmission of these viruses (6, 8). The occupational influenza vaccination programme for health and social care workers in the UK was introduced in 2003, and both groups have been prioritised for COVID-19 vaccination since its first roll-out late 2020. In 2024, the RSV vaccine programme was introduced in the UK, limited to those where the impact is greatest: older adults (75+) and pregnant women (to provide passive immunisation to infants). This is not currently recommended as an occupational vaccine (8).

Understanding the epidemiology of these viruses in HCWs over winter, including their impact on sick leave, and the potential role of seasonal vaccination in reducing this (9), is important for health service planning. Capturing infections in HCWs, particularly asymptomatic infections, may help understand nosocomial transmission dynamics and identify potential opportunities for intervention.

The SARS-CoV-2 Immunity and Reinfection Evaluation (SIREN) study is a prospective cohort study of HCWs in the UK, set up in June 2020, with regular SARS-CoV-2 polymerase-chain reaction (PCR) and antibody testing (10). Between November 2022 to March 2023, the SIREN Winter Pressures pilot study introduced multiplex testing into the cohort, testing for influenza and RSV alongside SARS-CoV-2 (11). The use of multiplex PCR allowed a better understanding of the dynamic of respiratory viruses during winter, however late deployment of multiplex PCR compounded by an unusually early influenza season, meant surveillance of the three viruses over winter 2022/23 was incomplete (12)(13).

Building on the pilot study, we aimed to measure the incidence of influenza, SARS-CoV-2 and RSV in UK HCWs over winter 2023/24, to characterise the symptom profiles, incidence of asymptomatic infections and the overall burden of these infections, and to describe sick leave by infection and vaccination status.

## METHODS

### Study design

A prospective cohort study, with participants undergoing follow-up between September 2023 and 31 March 2024 (10).

### Participants

Participants were HCWs aged 18 years and over recruited from secondary care settings across the UK. Participants were recruited into “SIREN 2.0” between August and September 2023 from the original SIREN cohort of 44,500 HCWs. Participants were included in this analysis if they completed both surveys and samples for PCR testing between 01 September 2023 and 31 March 2024.

### Data collection

Upon enrolment to the SIREN study, participants provided information on demographics, occupation, comorbidities and household composition via a baseline survey, conducted in 2020/21. Updated information was obtained at time of re-consenting to continue testing during 2023/24.

Participants completed a fortnightly survey to provide information on symptoms, and sickness absence. Influenza and COVID-19 vaccination data was self-reported via the fortnightly surveys and also obtained through linkage to national vaccination registries for completeness.

Participants self-administered nasal and throat swabs fortnightly at home and posted these to a centralised laboratory for nucleic acid amplification testing using a multiplex PCR assay for SARS-CoV-2, influenza A/B and RSV. Swabs for PCR testing were completed on a fixed fortnightly schedule regardless of symptoms. Positive samples for SARS-CoV-2, were whole genomic sequenced using SARS-CoV-2 ARTIC amplicon sequencing (14). Influenza and RSV positive samples were sequenced using bait-capture sequencing using a custom sequencing approach from the Welcome Sanger Institute using TWIST Respiratory Virus Research Panel (Ref to follow). Multiplex PCR test results, SARS-CoV-2 variant calls, influenza and RSV typing, symptoms and sickness absence data, and sociodemographic information were linked via a unique study ID.

### Outcomes

The co-primary outcomes were 1) PCR-positive test for either SARS-CoV-2, influenza, or RSV; and 2) the number of days taken off work due to reported symptoms.

### Variables

PCR samples that were positive for more than one virus were excluded from the analysis. Positive samples were de-duplicated to one sample per fortnight per participant, prioritising a positive sample over a negative sample in the same fortnight, and to one sample per infection episode. Infection episode was defined as one positive sample per participant within 90-days for SARS-CoV-2 and 30-days for influenza and RSV (15-18).

Three symptom categories were defined: influenza-like illness (ILI), respiratory symptoms and any symptoms. For this analysis, ILI was defined as fever and either cough, sore throat, shortness of breath, runny nose, headache, muscle ache or fatigue. The symptom onset date of any other symptom must occur within five days of onset of fever. For respiratory symptoms, participants had to report two or more of the following symptoms: respiratory (fever, cough, sore throat, shortness of breath, or runny nose) or other (headache, muscle ache, fatigue, joint pain, nausea, or diarrhoea). Where at least one of the symptoms must be respiratory and symptoms onset date of any respiratory or other symptom must occur within five days of onset of earliest respiratory symptom. Any symptoms were defined as participants reporting one of the following symptoms in the fortnightly survey: cough, fever, shortness of breath, sore throat, runny nose, headache, muscle aches, altered sense of smell or taste, fatigue, diarrhoea, nausea or vomiting, itchy red patches, rash, swollen glands, low mood, loss of concentration or brain fog and loss of appetite.

An infection was considered symptomatic if the participant had reported a symptom onset date within seven days either before or after the sample date for the first PCR positive result of the infection episode.

Vaccination in this analysis refers to the COVID-19 Autumn 2023 vaccine dose and the 2023/24 seasonal influenza vaccine between September 2023 and March 2024. Participants were considered vaccinated 14 days after receiving the COVID-19 Autumn 2023 dose or 2023/24 seasonal influenza vaccine.

Sickness absence was defined as self-reported absence and the number of days taken off work due to reported symptoms.

### Statistical analysis

Sociodemographic and occupational characteristics were described, overall and by infection (SARS-CoV-2, influenza and RSV). Positivity rates for each infection (SARS-CoV-2, influenza or RSV), with 95% confidence intervals, were described over time by fortnightly period. The proportion of positive SARS-CoV-2, influenza and RSV samples sequenced was calculated and trends described over time. Symptom type and duration, medical care attendance and sickness absence was compared by infection. Proportions were compared using Fisher exact test where applicable. The proportion of participants reporting symptoms by the three symptom definitions was calculated and trends described over time.

The number and proportion of participants who reported taking sick leave for their symptoms was calculated, regardless of a confirmed infection, by demographics and vaccination status, as well as the median and interquartile range (IQR) of the number of days taken.

Sick leave rate was calculated by fortnight, using the number of days taken off work divided by the number of days contributed to the analysis period, per 100 days, with 95% confidence interval.

Mixed-effects logistic regression was used to estimate the association between taking sick leave (yes/no) and vaccination status. As sick leave was reported as a total per fortnight and sick leave start date was not collected, data was transformed from fortnightly to daily reporting of sick leave, using symptom onset date as a proxy for the first date sick leave was taken. For each sick leave day, vaccination status was determined by if the participant had been vaccinated at least 14 days before the first day of sick leave. Sick leave taken between 0 to 13 days after vaccination was not included in analysis. For this model, vaccination status was defined as 1) not vaccinated for the COVID-19 Autumn 2023 dose or seasonal influenza 2023/24; 2) vaccinated for COVID-19 (Autumn 2023) and/or seasonal influenza 2023/24. To adjust for difference due to calendar time, month was included into the model, as well as socio-demographic factors (age group, gender, occupation, comorbidities). An additional sub-group analysis was conducted by restricting the model to only participants who had a SARS-CoV-2 or an influenza infection, and the corresponding vaccination (vs not being vaccinated), separately and had reported taking sick leave within the 14-day follow-up period.

## RESULTS

A total of 5,287 participants were included in the SIREN 2.0 analysis from 01 September 2023 to 31 March 2024 (Figure 1). Most participants were female (78·3%), of white ethnicity (85·7%), with a median age of 54 (IQR: 46-60) and the most common staff group was nursing (30·9%) (Table 1). Participants were highly vaccinated, with 66.0% receiving the COVID-19 Autumn 2023 vaccine dose, 74·8% receiving the 2023/24 seasonal influenza vaccine, and 61·6% receiving both.

**Figure 1:**
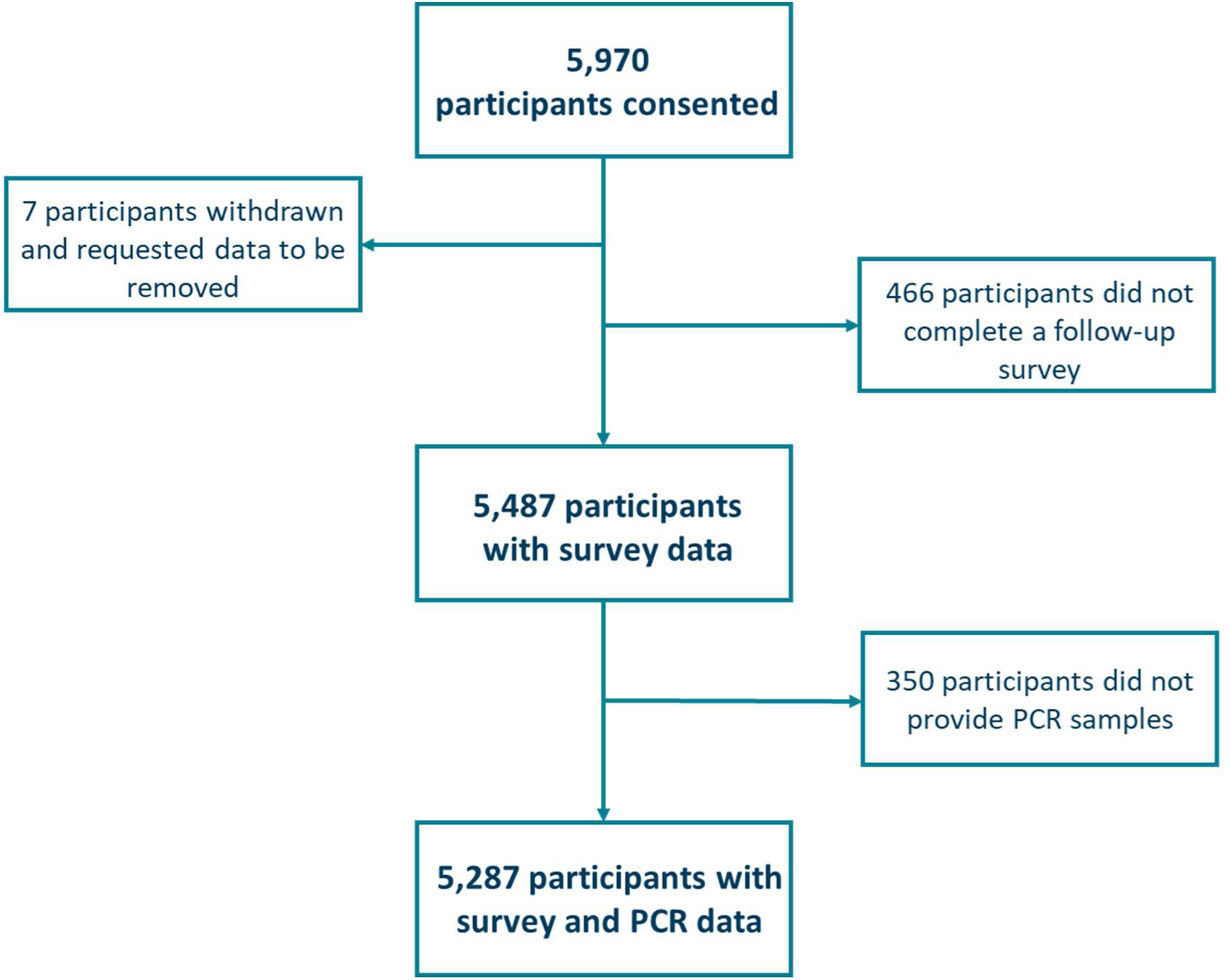
Flow diagram describing participants included in the SIREN Winter Pressure study.

**Table 1:** Demographics of participants included in the SIREN Winter Pressure 2023/24 study, overall and by infection (SARS-CoV-2, influenza and RSV)

| Characteristic | All participants<br>n (%) | SARS-CoV-2<br>positive<br>n (%) | Influenza<br>positive<br>n (%) | RSV<br>positive<br>n (%) |
| --- | --- | --- | --- | --- |
| **Total** | **5,287** | **1,150** | **278** | **384** |
| **Age group** |  |  |  |  |
| Under 35 | 202 (3.8) | 38 (3.3) | 8 (2.9) | 16 (4.2) |
| 35 to 44 | 831 (15.7) | 185 (16.1) | 54 (19.4) | 67 (17.4) |
| 45 to 54 | 1,738 (32.9) | 389 (33.8) | 96 (34.5) | 121 (31.5) |
| 55 to 64 | 2,104 (39.8) | 435 (37.8) | 98 (35.3) | 154 (40.1) |
| Over 65 | 412 (7.8) | 103 (9.0) | 22 (7.9) | 26 (6.8) |
| **Gender** |  |  |  |  |
| Female | 4,139 (78.3) | 909 (79.0) | 214 (77.0) | 309 (80.5) |
| Male | 1,144 (21.6) | 241 (21.0) | 64 (23.0) | 75 (19.5) |
| Non-binary | <5 | <5 | <5 | <5 |
| Prefer not to say | <5 | <5 | <5 | <5 |
| **Ethnicity** |  |  |  |  |
| White | 4,530 (85.7) | 998 (86.8) | 228 (82.0) | 328 (85.4) |
| Asian | 390 (7.4) | 74 (6.4) | 23 (8.3) | 27 (7.0) |
| Black | 168 (3.2) | 35 (3.0) | 13 (4.7) | 10 (2.6) |
| Mixed Race | 109 (2.1) | 23 (2.0) | 9 (3.2) | 12 (3.1) |
| Other Ethnic Group | 79 (1.5) | 18 (1.6) | 5 (1.8) | 6 (1.6) |
| Prefer not to say | 11 (0.2) | <5 | <5 | <5 |
| **Medical condition** |  |  |  |  |
| Chronic Respiratory conditions | 660 (12.5) | 152 (13.2) | 31 (11.2) | 64 (16.7) |
| Chronic Non-Respiratory conditions | 640 (12.1) | 139 (12.1) | 42 (15.1) | 42 (10.9) |
| Immunosuppression | 131 (2.5) | 26 (2.3) | 5 (1.8) | 6 (1.6) |
| No medical condition | 3,856 (72.9) | 833 (72.4) | 200 (71.9) | 272 (70.8) |
| **Household** |  |  |  |  |
| Lives alone | 674 (12.7) | 137 (11.9) | 36 (12.9) | 49 (12.8) |
| Lives with adults | 2,618 (49.5) | 556 (48.3) | 126 (45.3) | 186 (48.4) |
| Lives with children | 1,980 (37.5) | 455 (39.6) | 115 (41.4) | 149 (38.8) |
| Prefer not to say | 15 (0.3) | <5 | <5 | <5 |
| **Staff type** |  |  |  |  |
| Nursing | 1,632 (30.9) | 355 (30.9) | 83 (29.9) | 118 (30.7) |
| Administrative/Executive | 853 (16.1) | 175 (15.2) | 35 (12.6) | 64 (16.7) |
| Doctor | 753 (14.2) | 165 (14.3) | 53 (19.1) | 50 (13.0) |
| Healthcare Assistant | 314 (5.9) | 64 (5.6) | 18 (6.5) | 25 (6.5) |
| Healthcare Scientist | 378 (7.1) | 87 (7.6) | 17 (6.1) | 27 (7.0) |
| Student | 128 (2.4) | 33 (2.9) | 6 (2.2) | 7 (1.8) |
| Therapist <sup>a</sup> | 245 (4.6) | 51 (4.4) | 10 (3.6) | 25 (6.5) |
| Midwife | 190 (3.6) | 46 (4.0) | 9 (3.2) | 14 (3.6) |
| Pharmacist | 141 (2.7) | 30 (2.6) | 7 (2.5) | 9 (2.3) |
| Estates/Porters/Security | 119 (2.3) | 23 (2.0) | 8 (2.9) | 6 (1.6) |
| Other | 534 (10.1) | 121 (10.5) | 32 (11.5) | 39 (10.2) |
| **Occupation setting** |  |  |  |  |
| Home Working | 382 (7.2) | 79 (6.9) | 20 (7.2) | 28 (7.3) |
| Office | 526 (9.9) | 135 (11.7) | 24 (8.6) | 43 (11.2) |
| Outpatient | 1,851 (35.0) | 378 (32.9) | 104 (37.4) | 125 (32.6) |
| Inpatient Wards | 310 (5.9) | 60 (5.2) | 14 (5.0) | 23 (6.0) |
| Patient facing (non-clinical) | 268 (5.1) | 65 (5.7) | 17 (6.1) | 17 (4.4) |
| Intensive Care | 625 (11.8) | 125 (10.9) | 37 (13.3) | 49 (12.8) |
| Theatres | 57 (1.1) | 11 (1.0) | 5 (1.8) | <5 |
| Ambulance/Emergency Department | 157 (3.0) | 27 (2.3) | 13 (4.7) | 14 (3.6) |
| Maternity/Labour Ward | 166 (3.1) | 46 (4.0) | 7 (2.5) | 10 (2.6) |
| Other | 945 (17.9) | 224 (19.5) | 37 (13.3) | 74 (19.3) |
| **Patient contact** |  |  |  |  |
| Yes | 4,291 (81.2) | 933 (81.1) | 237 (85.3) | 299 (77.9) |
| No | 996 (18.8) | 217 (18.9) | 41 (14.7) | 85 (22.1) |

*Note: numbers under five have been suppressed (<5). Participants who were infected with more than one virus over the analysis period are included in each of the applicable columns.*
<sup>a</sup> *Therapist includes: Physiotherapist, Occupational Therapist, and Speech and Language Therapists*

### SARS-CoV-2, influenza A/B and RSV infection trends over winter 2023/24

Between September 2023 and March 2024, there were 1,828 infections among 1,659 participants (1,161 SARS-CoV-2; 387 RSV; 280 influenza infections). Over the observation period 21·8% of participants had at least one SARS-CoV-2 infection, 7·3% had at least one RSV infection and 5·3% had at least one influenza infection.

There were 149 participants who were infected with more than one virus during the analysis period. Of these, 68 participants were infected with SARS-CoV-2 and RSV, 54 with SARS-CoV-2 and influenza, 23 with influenza and RSV, and four participants were infected with all three viruses. The median time between infections was 55 days (IQR: 29-84 days). Additionally, during this period, 11 participants had two SARS-CoV-2 infections >90 days apart, three participants with two RSV infections >30 days apart, and two participants with two influenza infections >30 days apart. The median time between infections for each virus was 108 days for SARS-CoV-2 (IQR: 97-128), 40 days for RSV (IQR: 31-168), and 56·5 days for influenza (IQR: 43-70).

Fortnightly infection rates varied, with SARS-CoV-2 positivity peaking in both September and December 2023 (4·0% and 4·3%, respectively). Influenza positivity peaked in mid-December 2023 (1·2%) and maintained a similar positivity rate until mid-February 2024. RSV peaked in early December (1·9%), with positivity decreasing to low levels from early January 2024 (Figure 2).

**Figure 2:**
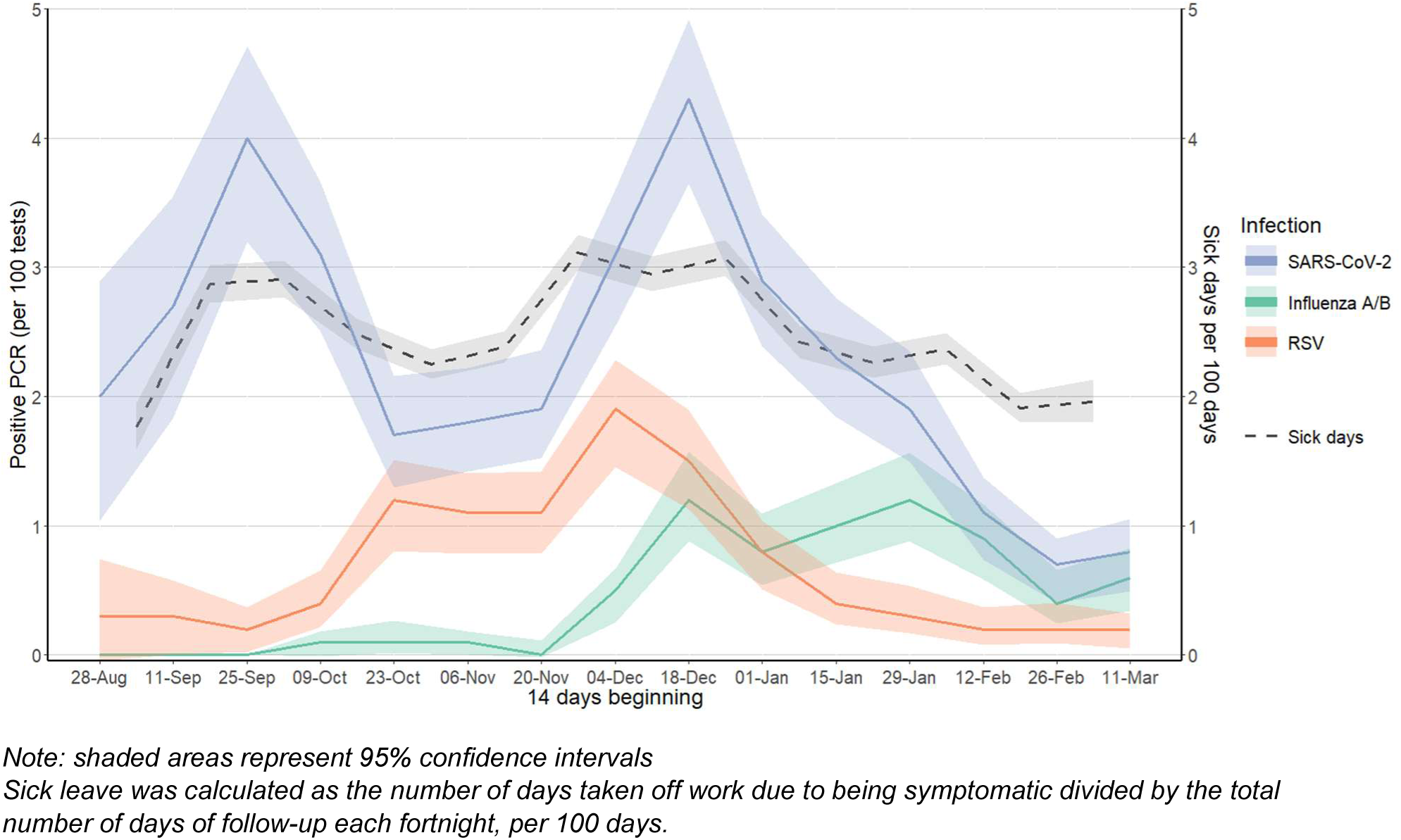
PCR positivity (per 100 tests) by infection (SARS-CoV-2, influenza and RSV) and sick leave rate (per 100 days follow-up), within SIREN Winter Pressure 2023/24 study.

16·8% (587/3,489) of vaccinated participants received the COVID-19 vaccine by 25 September 2023 (the first peak of SARS-CoV-2 positivity) and 98·6% by the 18 December 2023 (the second peak of SARS-CoV-2 positivity). 98·8% (3,905/3,954) of vaccinated participants received the influenza vaccine by 18 December 2023 (the peak of influenza positivity).

Genomics derived SARS-CoV-2 variant data was available for 758 (65·3%) positive SARS-CoV-2 samples. During the first peak of SARS-CoV-2 (between 28 August 2023 and 22 October 2023), 31·2% (48/154) of sequenced samples were XBB variant. In the second peak (between 20 November 2023 and 27 January 2024), 63·4% (248/391) of sequenced samples were JN.1 variant. Genomic sequencing of for RSV samples were successful for, 56·8% (220/387) were typed, of which 58·2% were type A and 41·8% type B. For influenza, 77·9% (218/280) of samples were sequenced, of which 58·8% were H1N1, 36·2% were H3N2, and 5·1% were influenza B (Supplementary Table 1).

### Symptom profile and sick leave taken by infection (SARS-CoV-2, influenza A/B and RSV)

The proportion of asymptomatic infections was highest for RSV (30·7%) compared with 19·0% for influenza and 19·8% for SARS-CoV-2 (RSV vs influenza or SARS-CoV-2, p≤0·001). Symptom profiles were broadly similar across the three infections, with runny nose, sore throat and headache the most frequent symptoms reported. Symptoms reported for SARS-CoV-2 and influenza were more similar compared to RSV, except for cough and fever, which was more frequently reported for influenza infections (p≤0·01) (Figure 3). The median duration of symptoms reported was 7 days for all three viruses (Table 2). Symptomatic infections by influenza type were similar and for SARS-CoV-2 and RSV differed between variants/type (Supplementary Table 1).

**Figure 3:**
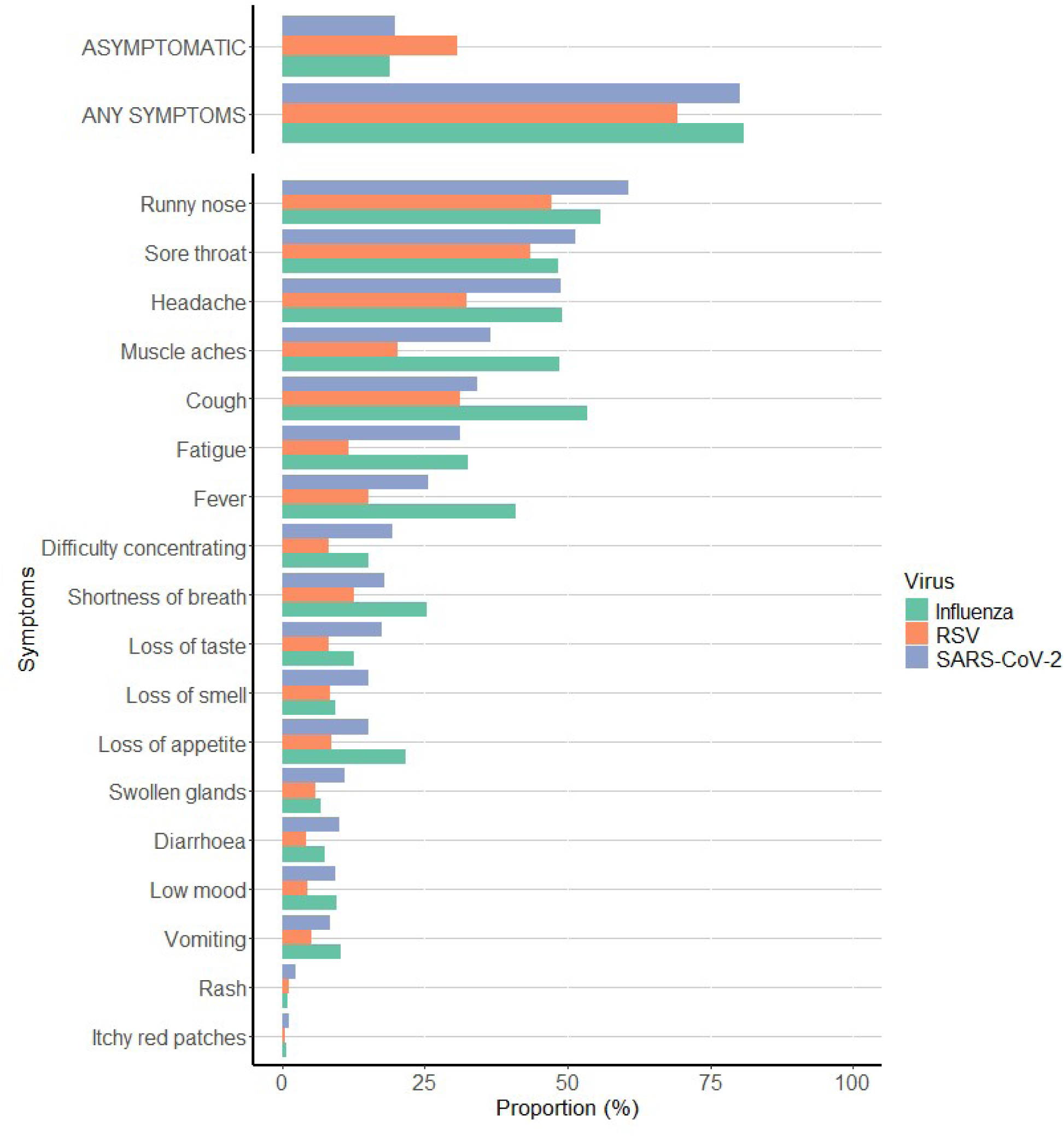
Proportion of participants reporting symptoms by infection (SARS-CoV-2, influenza and RSV), within SIREN Winter Pressure 2023/24 study.

**Table 2:** Number and proportion of symptomatic PCR positive samples by infection^a^ (SARS-CoV-2, influenza and RSV)

|  | **SARS-CoV-2**<br>**n (%)** | **Influenza**<br>**n (%)** | **RSV**<br>**n (%)** | *p-value*<br>*(SARS-CoV-2*<br>*vs influenza)* | *p-value*<br>*(SARS-*<br>*CoV-2*<br>*vs RSV)* | *p-value*<br>*(RSV vs*<br>*influenza)* |
| --- | --- | --- | --- | --- | --- | --- |
| **Total** | **1,161** | **281** | **387** |  |  |  |
| **Symptoms** |  |  |  | *0.798* | *<0.001* | *0.001* |
| Any symptoms | 893 (80.2) | 218 (81.0) | 257 (69.3) |  |  |  |
| Asymptomatic | 221 (19.8) | 51 (19.0) | 114 (30.7) |  |  |  |
| **Symptoms type** |  |  |  |  |  |  |
| Loss of smell | 169 (15.2) | 25 (9.3) | 31 (8.4) | *0.014* | *<0.001* | *0.674* |
| Cough | 380 (34.1) | 144 (53.5) | 116 (31.3) | *<0.001* | *0.341* | *<0.001* |
| Diarrhoea | 111 (10.0) | 20 (7.4) | 16 (4.3) | *0.246* | *<0.001* | *0.117* |
| Loss of taste | 195 (17.5) | 34 (12.6) | 30 (8.1) | *0.055* | *<0.001* | *0.063* |
| Fatigue | 348 (31.2) | 88 (32.7) | 43 (11.6) | *0.661* | *<0.001* | *<0.001* |
| Fever | 285 (25.6) | 110 (40.9) | 56 (15.1) | *<0.001* | *<0.001* | *<0.001* |
| Swollen glands | 123 (11.0) | 18 (6.7) | 22 (5.9) | *0.033* | *0.003* | *0.742* |
| Headache | 544 (48.8) | 132 (49.1) | 120 (32.3) | *0.946* | *<0.001* | *<0.001* |
| Muscle aches | 408 (36.6) | 131 (48.7) | 75 (20.2) | *<0.001* | *<0.001* | *<0.001* |
| Runny nose | 675 (60.6) | 150 (55.8) | 175 (47.2) | *0.166* | *<0.001* | *0.037* |
| Shortness of breath | 200 (18.0) | 68 (25.3) | 47 (12.7) | *0.008* | *0.019* | *<0.001* |
| Sore throat | 573 (51.4) | 130 (48.3) | 161 (43.4) | *0.377* | *0.008* | *0.228* |
| Vomiting | 94 (8.4) | 28 (10.4) | 19 (5.1) | *0.337* | *0.041* | *0.014* |
| **Duration of symptoms** |  |  |  | *0.301* | *0.857* | *0.310* |
| Median (days [IQR]) | 7 [4-12] | 7 [3-12.5] | 7 [4-12] |  |  |  |
| **Sick leave** |  |  |  | *0.880* | *<0.001* | *0.003* |
| Taken | 414 (46.6) | 100 (45.9) | 83 (32.5) |  |  |  |
| Not taken | 475 (53.4) | 118 (54.1) | 172 (67.5) |  |  |  |
| **Sick leave taken (days)** |  |  |  |  |  |  |
| Median [IQR] | 3 [2-5] | 3 [2-5] | 3 [1-5] |  |  |  |
| **Sought medical care** |  |  |  | *<0.001* | *0.002* | *0.119* |
| Yes - admitted to hospital | 4 (0.5) | 3 (1.4) | 1 (0.4) |  |  |  |
| Yes - A&E but not admitted | 2 (0.2) | 6 (2.8) | 2 (0.8) |  |  |  |
| Yes - primary care | 55 (6.2) | 36 (16.6) | 33 (12.9) |  |  |  |
| No | 826 (93.1) | 172 (79.3) | 219 (85.9) |  |  |  |

<sup>a</sup> 149 participants had multiple infections over the analysis period – 68 with a SARS-CoV-2 and an RSV infection, 54 with a SARS-CoV-2 and an influenza infection, 23 with an RSV and an influenza infections, and four participants had all three infections over the analysis period
Note: p-values calculated using Fisher's exact test.

Overall, 7·2% (137/1,659) of participants reported seeking medical care (including visiting GP practice (120), accident and emergency department (10) or being admitted to hospital (3)) for their symptoms following an infection. Participants with SARS-CoV-2 were less likely to seek medical care for their symptoms (6·9%) than those with RSV (14·1%) or influenza (20·8%). Those with influenza were most likely to attend accident and emergency department or to be admitted to hospital (4·2%, vs SARS-CoV-2: 0·7% and RSV: 1·2%) (Table 2).

34·6% (574/1,659) of participants who tested positive for at least one of the three viruses reported taking sick leave, resulting in a total 2,376 days of sick leave taken (1,705 days was taken for COVID-19 infections, 361 for influenza, and 310 for RSV).

Comparing the proportion of participants reporting sick leave (taking vs not taking) by infection, participants with an RSV infection reported taking less sick leave in general, compared to the other infections (32·5% vs influenza: 45·9% and SARS-CoV-2: 46·6%, p≤0·003), although the median number of sick days was similar between infections (three days).

For those with a SARS-CoV-2 or influenza infection, the proportion of participants taking sick leave did not differ by vaccination status (SARS-CoV-2 not vaccinated 50·4% vs vaccinated 42·6%, p=0·029; influenza not vaccinated 46·3% vs vaccinated 45·7%; p>0·999). Participants who were not vaccinated at the time of their SARS-CoV-2 infection took a median of four days sick leave (IQR: 2-5), with those who were vaccinated taking a median of three days (IQR: 2-5). No difference was seen in the number of days taken for influenza infections by vaccination status (median three days).

### Symptoms trends and sick leave by demographics and vaccination status

85·9% (4,540/5,287) participants reported symptoms during the analysis period, regardless of a known infection. Trends by symptom category (any symptoms, respiratory symptoms and ILI symptoms) were similar over time. This trend was also similar to the SARS-CoV-2 PCR positivity, with a peak in September and December (Figure 2 and Figure 4).

**Figure 4:**
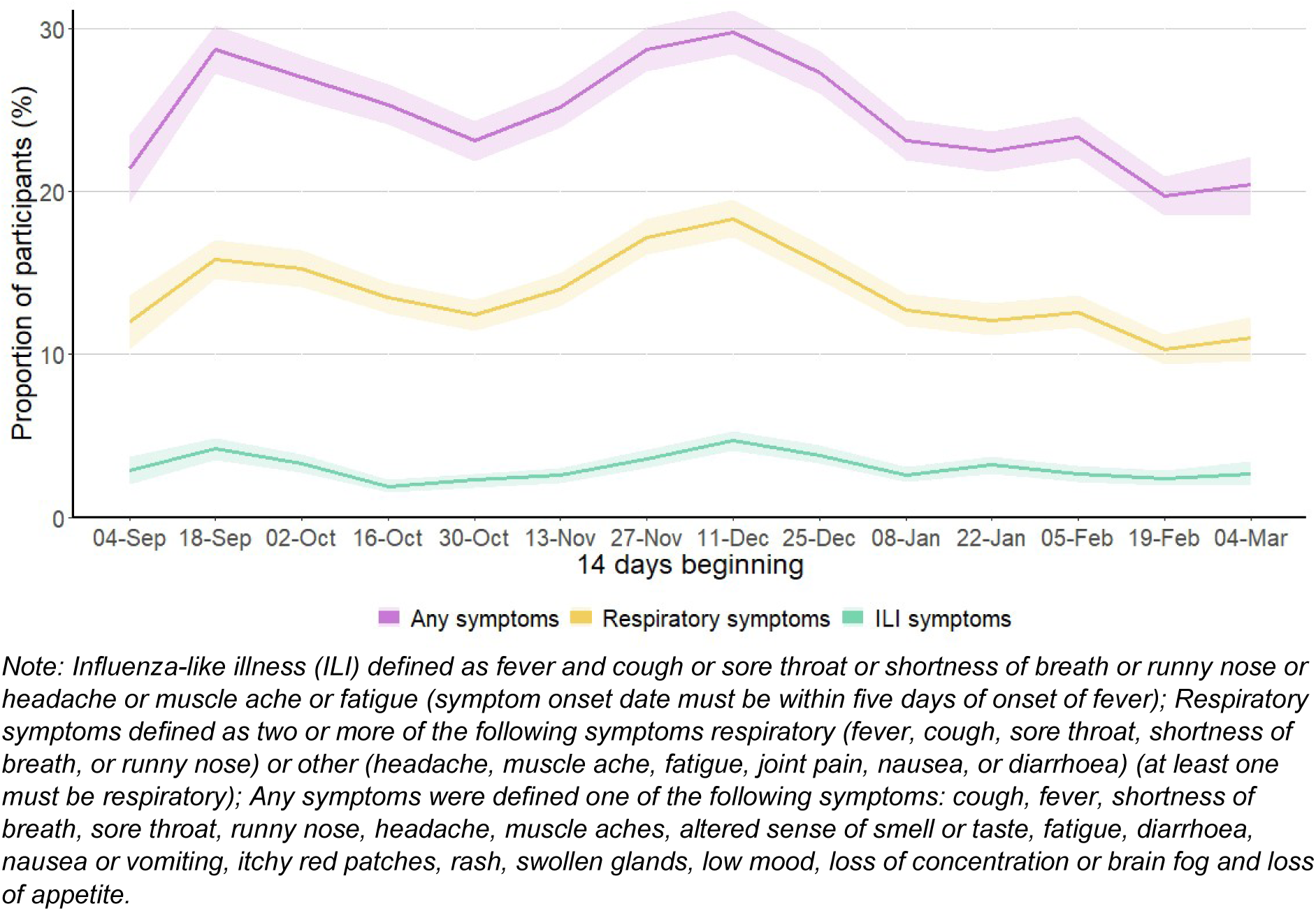
Proportion of participants reporting symptoms over time by symptom category (any symptom, respiratory symptom and influenza-like illness)

Overall, regardless of a known infection, 35·8% (1,892/5,287) of participants reported taking sick leave, resulting in a total of 10,168 days of sick leave taken over the analysis period and a median of five days taken per person (range 1-98 days).

The proportion of participants taking sick leave and the number of days of sick leave were similar between demographic, occupation, and vaccination status (Table 3). The only notable differences observed were among those with a reported medical condition (no medical condition: median of three days of sick leave taken compared to five days among immunosuppressed participants). Those who worked in theatres also took more days of sick leave compared those who work in an outpatient, inpatient ward, intensive care, ambulance/emergency department or other occupation setting (median of seven days versus three days, respectively).

**Table 3:** Number of participants reporting sick leave and the median and interquartile range (IQR) of days taken by demographics and vaccination status.

| Demographics / Vaccination status | Total participants reporting on sick leave (Yes/No) | Total taking sick leave (n/%) | Median [IRQ] days of sick leave taken |
| --- | --- | --- | --- |
| **Age group** |  |  |  |
| Under 35 | 173 | 77 (44.5) | 4 [2-8] |
| 35 to 44 | 741 | 347 (46.8) | 3 [2-6] |
| 45 to 54 | 1494 | 658 (44.0) | 3 [2-6] |
| 55 to 64 | 1778 | 704 (39.6) | 4 [2-7] |
| Over 65 | 319 | 106 (33.2) | 4 [2-7] |
| **Gender** |  |  |  |
| Female | 3558 | 1548 (43.5) | 4 [2-7] |
| Male | 944 | 342 (36.2) | 4 [2-7] |
| Non-binary | <5 | <5 | 2.5 [2-3] |
| **Ethnicity** |  |  |  |
| White | 3896 | 1633 (41.9) | 3 [2-7] |
| Asian | 311 | 125 (40.2) | 3 [2-6] |
| Black | 128 | 57 (44.5) | 5 [2-8] |
| Mixed Race | 99 | 41 (41.4) | 3 [2-5] |
| Other Ethnic Group | 62 | 29 (46.8) | 5 [2-7] |
| Prefer not to say | 10 | 7 (70.0) | 5 [3-8] |
| **Medical group** |  |  |  |
| No medical condition | 3265 | 1313 (40.2) | 3 [2-6] |
| Chronic respiratory conditions | 571 | 283 (49.6) | 4 [2-7] |
| Chronic non-respiratory conditions | 556 | 249 (44.8) | 4 [2-7] |
| Immunosuppression | 114 | 47 (41.2) | 5 [2-10] |
| **Household** |  |  |  |
| Lives with adults | 2207 | 895 (40.6) | 4 [2-7] |
| Lives with children | 1726 | 748 (43.3) | 3 [2-6] |
| Lives alone | 559 | 244 (43.6) | 3 [2-6] |
| **Staff type** |  |  |  |
| Nursing | 1398 | 583 (41.7) | 3 [2-7] |
| Administrative/Executive | 697 | 276 (39.6) | 3 [2-6] |
| Doctor | 667 | 219 (32.8) | 3 [1-5] |
| Healthcare Scientist | 324 | 154 (47.5) | 4 [2-7] |
| Healthcare Assistant | 250 | 129 (51.6) | 5 [2-9] |
| Therapist <sup>a</sup> | 219 | 110 (50.2) | 3 [2-6] |
| Midwife | 159 | 62 (39.0) | 4 [2-7] |
| Pharmacist | 128 | 48 (37.5) | 3 [2-7] |
| Student | 108 | 48 (44.4) | 4.5 [2.5-6] |
| Estates/Porters/Security | 89 | 41 (46.1) | 5 [3-8] |
| Other | 467 | 222 (47.5) | 4 [2-6] |
| **Occupation setting** |  |  |  |
| Outpatient | 1579 | 650 (41.2) | 3 [2-7] |
| Inpatient Wards | 270 | 125 (46.3) | 3 [2-7] |
| Intensive Care | 528 | 234 (44.3) | 3 [2-7] |
| Patient facing (non-clinical) | 234 | 95 (40.6) | 4 [2-7] |
| Maternity/Labour Ward | 146 | 62 (42.5) | 4 [2-5] |
| Home Working | 298 | 118 (39.6) | 4 [2-7] |
| Ambulance/Emergency Department | 130 | 53 (40.8) | 3 [1-5] |
| Office | 447 | 184 (41.2) | 4 [2-6] |
| Theatres | 42 | 19 (45.2) | 7 [3-10] |
| Other | 832 | 352 (42.3) | 3 [2-7] |
| **Patient contact** |  |  |  |
| Yes | 3659 | 1542 (42.1) | 4 [2-7] |
| No | 847 | 350 (41.3) | 3 [2-6] |
| **Vaccination status** |  |  |  |
| Did not receive the COVID-19 Autumn 2023 dose or the 2023/24 seasonal influenza vaccine | - | - | 3 [2-6] |
| Received both the COVID-19 Autumn 2023 dose and the 2023/24 seasonal influenza vaccine | - | - | 3 [1-6] |
| Received the COVID-19 Autumn 2023 dose only | - | - | 3 [2-7] |
| Received the 2023/24 seasonal influenza vaccine only | - | - | 3 [2-6] |

*Note: Vaccination status: participants could be included in multiple categories, as participants could move from an unvaccinated (COVID-19 Autumn 2023 dose and seasonal Influenza) to one or more of the vaccinated categories; 2,870 participants provided time to the unvaccinated status with 632 (22.0%) participants taking sick leave; 874/2,746 (31.8%) for participants who received both vaccinations, 92/302 (30.5%) for participants who received the COVID-19 Autumn 2023 dose only and 245/783 (31.3%) for participants who received the 2023/24 seasonal influenza vaccine only; numbers under five have been suppressed (<5)*
<sup>a</sup> *Therapist includes: Physiotherapist, Occupational Therapist, and Speech and Language Therapists*

The logistic regression model did not indicate any difference in the likelihood of taking sick leave by vaccination status (adjusted odd ratios (aOR) 0·98; 95% CI 0·87, 1·10) (Table 4). No differences were observed in the SARS-CoV-2 infection only model (Supplementary Table 2) and the influenza model results were inconclusive, due to lower number of infections.

**Table 4:** Factors associated with taking sick leave.

|  | Unadjusted |  |  | Adjusted |  |  |
| --- | --- | --- | --- | --- | --- | --- |
|  | Odds ratio | 95% Confidence Interval | p-value | Odds ratio | 95% Confidence Interval | p-value |
| **Vaccination status** |  |  |  |  |  |  |
| Not vaccinated | Ref |  |  | Ref |  |  |
| Vaccinated | 1.18 | 1.07, 1.29 | <0.00 | 0.98 | 0.87, 1.10 | 0.69 |
| **Month of sick leave** |  |  |  |  |  |  |
| September | Ref |  |  | Ref |  |  |
| October | 1.10 | 0.97, 1.25 | 0.12 | 1.24 | 1.06, 1.46 | 0.01 |
| November | 0.89 | 0.78, 1.01 | 0.07 | 1.07 | 0.90, 1.28 | 0.44 |
| December | 1.36 | 1.21, 1.53 | <0.00 | 1.52 | 1.27, 1.81 | <0.00 |
| January | 1.34 | 1.19, 1.52 | <0.00 | 1.38 | 1.15, 1.65 | <0.00 |
| February | 1.04 | 0.91, 1.18 | 0.61 | 1.27 | 1.05, 1.53 | 0.01 |
| March | 1.77 | 1.47, 2.14 | <0.00 | 1.65 | 1.33, 2.05 | <0.00 |
| **Age group** |  |  |  |  |  |  |
| Under 25 | Ref |  |  | Ref |  |  |
| 25 to 34 | 0.66 | 0.21, 2.09 | 0.48 | 0.41 | 0.18, 0.9 | 0.03 |
| 35 to 44 | 0.53 | 0.17, 1.64 | 0.27 | 0.38 | 0.17, 0.83 | 0.02 |
| 45 to 54 | 0.59 | 0.19, 1.81 | 0.36 | 0.43 | 0.20, 0.95 | 0.04 |
| 55 to 64 | 0.52 | 0.17, 0.62 | 0.26 | 0.46 | 0.21, 1.01 | 0.05 |
| Over 65 | 0.39 | 0.11, 1.36 | 0.14 | 0.52 | 0.22, 1.26 | 0.15 |
| **Gender** |  |  |  |  |  |  |
| Male | Ref |  |  | Ref |  |  |
| Female | 1.3 | 1.09, 1.62 | <0.00 | 0.87 | 0.75, 1.00 | 0.06 |
| Non-binary | 0.32 | 0.25, 42.61 | 0.37 | 0.92 | 0.18, 4.71 | 0.92 |
| **Staff group** |  |  |  |  |  |  |
| Administrative | Ref |  |  | Ref |  |  |
| Doctor | 0.60 | 0.43, 0.76 | <0.00 | 0.75 | 0.61, 0.93 | 0.01 |
| Nursing | 0.90 | 0.78, 1.26 | 0.92 | 1.00 | 0.84, 1.18 | 0.96 |
| Healthcare Assistant | 1.90 | 1.30, 2.71 | <0.00 | 1.18 | 0.92, 1.5 | 0.19 |
| Midwife | 0.94 | 0.60, 1.49 | 0.80 | 1.01 | 0.73, 1.38 | 0.97 |
| Healthcare Scientist | 1.40 | 0.99, 1.95 | 0.06 | 1.00 | 0.8, 1.26 | 0.99 |
| Pharmacist | 0.93 | 0.55, 1.57 | 0.77 | 1.00 | 0.7, 1.44 | 1.00 |
| Therapist<sup>a</sup> | 1.42 | 0.97, 2.07 | 0.07 | 0.99 | 0.77, 1.28 | 0.94 |
| Student | 1.38 | 0.87, 2.22 | 0.17 | 0.91 | 0.67, 1.25 | 0.57 |
| Estates/Porters | 1.21 | 0.65, 2.24 | 0.55 | 0.97 | 0.63, 1.49 | 0.89 |
| Other | 1.32 | 0.99, 1.77 | 0.06 | 0.99 | 0.81, 1.21 | 0.91 |
| **Medical group** |  |  |  |  |  |  |
| No medial conditions | Ref |  |  | Ref |  |  |
| Immunosuppression | 1.20 | 0.77, 2.02 | 0.38 | 1.55 | 1.12, 2.16 | 0.01 |
| Chronic respiratory conditions | 1.60 | 1.32, 2.05 | <0.00 | 1.17 | 1.02, 1.36 | 0.03 |
| Chronic non-respiratory conditions | 1.30 | 1.05, 1.66 | 0.02 | 1.02 | 0.87, 1.19 | 0.81 |

*Note: Not vaccinated defined as not receiving the COVID-19 Autumn 2023 dose or the 2023/24 seasonal influenza vaccine; Vaccinated as defined as receiving the COVID-19 Autumn 2023 dose and/or the 2023/24 seasonal influenza vaccine for at least 14 days*
<sup>a</sup> *Therapist includes: Physiotherapist, Occupational Therapist, and Speech and Language Therapists*

## DISCUSSION

Over winter 2023/24, 21% of UK healthcare workers participating in the SIREN study had a COVID-19 infection, 7% had an RSV infection and 5% had an influenza. 36% of participants took sick leave regardless of a known infection, with a combined total 10,168 days of sick leave taken over the analysis period. All three infections peaked in mid-December, compounding the impact on the health service, and underscoring the importance of combined monitoring of these viruses over winter. We observed differences in sickness absence by virus but did not find evidence that demographics, occupational setting, clinical history or seasonal vaccination majorly impacted sickness absence.

National surveillance data from winter 2023/24 showed similar trends to the SIREN data, including the double peak in SARS-CoV-2 driven by two distinct variants, and influenza peaking in mid-December 2023. Compared to 2022/23 winter season, influenza had a lower but more prolonged circulation, and RSV increased earlier (late October), but also peaking early/mid-December (19)(20). SIREN ILI trends were also consistent with national surveillance systems in the UK, including syndromic surveillance and the community based participatory FluSurvey, which reported an increase in GP in-hours consultations for ILI and ILI incidence starting in late-November and peaking in mid-December (19).

The similar clinical presentation of the three viral infections highlights the challenges to make an aetiological diagnosis from symptoms alone and the importance of multiplex testing. Asymptomatic infections were more common among RSV infections (30%), however, around 20% of SARS-CoV-2 and influenza infections were also asymptomatic. Rates of asymptomatic infection for COVID-19 have been well documented and are consistent with our findings (21), whereas rates of asymptomatic influenza are less well described with a large range of estimates depending on testing methodology (22-25), mostly from outbreak studies or serology. For RSV, most previous research has focused on the extremes of age, and, therefore, the asymptomatic fraction in a working aged population is largely unknown (26).

Our findings demonstrate that seasonal respiratory illness drives considerable staff sickness absence, particularly over short time windows, creating acute pressures on staffing, with considerable cost implications for healthcare organisations. For the 2023/24 winter season, since the much higher incidence of SARS-CoV-2 compared to influenza and RSV in our cohort, SARS-CoV-2 was the main contributor to sickness absence.

Given the scale of illness and sick leave associated with respiratory viruses over winter, interventions to protect the healthcare workforce from respiratory infections are important from an employer, occupational health and patient perspective. Seasonal influenza vaccination and COVID-19 Autumn 2023 vaccinations for NHS staff were both deployed as key interventions over winter 2023/24. We have analysed the effectiveness of both vaccines against infection and symptomatic infection in separate papers, and demonstrated that both vaccines did protect against infection, although the results were relatively modest (27), aligned with results from other HCW cohort studies (28). However, we did not find any evidence that seasonal vaccination reduced sick leave in this analysis.

A key strength of our study is the study design, with fortnightly multiplex molecular testing at scale combined with symptoms reporting throughout winter, ensuring systemic detections of infections throughout this period and providing a unique opportunity to characterise those infections, including the asymptomatic fraction. Having healthcare workers as a study population is an opportunity to reflect on infection trends of respiratory pathogens in a healthy, working age population, which is rarely tested in most settings. This is particularly relevant considering there is a gap of knowledge on the impact of RSV infections in this population, given the current evidence is focused on infants and older adults. By utilising a centralised approach, testing was delivered more efficiently within the study when compared to a pilot performed in winter 2022/23 (29), which resulted in higher-quality data and increased participants retention.

Our study has some limitations. Our multiplex PCR test was restricted to SARS-CoV-2, influenza and RSV and would not capture other pathogens that can cause respiratory illness. This highlights the value of other laboratory methods for surveillance, such as metagenomics which is currently being explored within our study. Our fortnightly testing interval was designed for COVID-19 and may have missed some influenza and RSV cases given the shorter duration of shedding (6, 7). Measuring sick leave in healthcare workers is complex, considering different working patterns, roles and settings. In this analysis, the proportion of participants taking sick leave and median were used to describe the number of days of sick leave. However, in the current phase of the study, more detailed questions regarding scheduled days at work and days taken off sick have been recorded, so that the rate of sick leave can be calculated. Although the model was unable to detect an association between vaccination and sick leave, further work to explore alternative statistical modelling approaches may be warranted.

## Conclusion

The burden of respiratory illness in our healthcare cohort over winter 2023/24 was substantial, with over one-third taking sickness absence for respiratory illness. SARS-CoV-2 had higher infection rates than influenza and RSV combined. All three viruses peaked simultaneously, compounding the impact on the health system. Workforce resilience over winter remains an important priority, highlighting the importance of effective interventions to reduce the burden of these viruses.

## Supporting information

Respiratory Virus and Microbiome Initiative

SIREN study group

## Data Availability

Anonymised data will be made available for secondary analysis to trusted researchers upon reasonable request.

---

**Supplementary Table 1:** Proportion of infections by variant/type and by symptomatic infections.

| Infection | Variant / type<br>n (%) | Symptomatic infection<br>n (%) <sup>a</sup> |
| --- | --- | --- |
| **SARS-CoV-2** |  |  |
| JN.1 | 365 (48.2) | 302 (85.8) |
| XBB | 60 (7.9) | 51 (85.0) |
| EG.5.1 | 41 (5.4) | 35 (94.6) |
| BA.2.86 | 34 (4.5) | 26 (76.5) |
| HV.1 | 27 (3.6) | 22 (84.6) |
| JN.2 | 21 (2.8) | 17 (85.0) |
| JG.3 | 20 (2.6) | 16 (80.0) |
| JD.1.1 | 18 (2.4) | 16 (94.1) |
| FL.1.5 | 17 (2.2) | 14 (82.4) |
| HK.3 | 16 (2.1) | 13 (81.3) |
| Other | 139 (18.3) | 118 (87.4) |
| **Influenza** |  |  |
| H1N1 | 127 (58.3) | 108 (88.5) |
| H3N2 | 79 (36.2) | 67 (88.2) |
| B | 11 (5.1) | 8 (80.0) |
| **RSV** |  |  |
| Type A | 128 (58.2) | 93 (74.4) |
| Type B | 92 (41.8) | 73 (84.9) |

<sup>a</sup> Where symptom data was reported

**Supplementary Table 2:** Estimates (odds ratios) investigating associations between sick leave and vaccination status among participants with a confirmed SARS-CoV-2 infection.

|  | Unadjusted |  |  | Adjusted |  |  |
| --- | --- | --- | --- | --- | --- | --- |
|  | Odds ratio | 95% Confidence Interval | p-value | Odds ratio | 95% Confidence Interval | p-value |
| **Vaccination status** |  |  |  |  |  |  |
| Not vaccinated | Ref |  |  | Ref |  |  |
| Vaccinated | 0.95 | 0.69, 1.32 | 0.77 | 0.95 | 0.66, 1.38 | 0.79 |
| **Month of sick leave** |  |  |  |  |  |  |
| September | Ref |  |  | Ref |  |  |
| October | 1.56 | 0.84, 2.89 | 0.16 | 1.64 | 0.88, 3.07 | 0.12 |
| November | 1.29 | 0.66, 2.51 | 0.45 | 1.32 | 0.66, 2.64 | 0.44 |
| December | 1.29 | 0.68, 2.44 | 0.43 | 1.37 | 0.71, 2.67 | 0.35 |
| January | 1.13 | 0.59, 2.19 | 0.71 | 1.14 | 0.58, 2.27 | 0.70 |
| February | 0.68 | 0.33, 1.42 | 0.31 | 0.69 | 0.32, 1.48 | 0.34 |
| March | 0.36 | 0.11, 1.19 | 0.09 | 0.30 | 0.09, 1.01 | 0.05 |
| **Age group** |  |  |  |  |  |  |
| Under 25 | Ref |  |  | Ref |  |  |
| 25 to 34 | 0.75 | 0.05, 10.43 | 0.83 | 1.02 | 0.07, 14.61 | 0.99 |
| 35 to 44 | 0.60 | 0.05, 7.89 | 0.70 | 1.05 | 0.08, 14.57 | 0.97 |
| 45 to 54 | 0.64 | 0.05, 8.31 | 0.73 | 1.05 | 0.08, 14.54 | 0.97 |
| 55 to 64 | 0.68 | 0.05, 8.92 | 0.77 | 1.13 | 0.08, 15.80 | 0.93 |
| Over 65 | 0.69 | 0.04, 10.86 | 0.79 | 1.02 | 0.06, 16.98 | 0.99 |
| **Gender** |  |  |  |  |  |  |
| Male | Ref |  |  | Ref |  |  |
| Female | 0.75 | 0.51, 1.12 | 0.16 | 0.64 | 0.41, 1.00 | 0.05 |
| **Staff group** |  |  |  |  |  |  |
| Administrative | Ref |  |  | Ref |  |  |
| Doctor | 0.89 | 0.47, 1.69 | 0.72 | 0.70 | 0.35, 1.38 | 0.31 |
| Nursing | 0.90 | 0.53, 1.52 | 0.69 | 0.92 | 0.54, 1.58 | 0.77 |
| Healthcare Assistant | 1.00 | 0.44, 2.27 | 1.00 | 1.17 | 0.50, 2.73 | 0.71 |
| Midwife | 0.89 | 0.34, 2.32 | 0.81 | 0.81 | 0.30, 2.17 | 0.67 |
| Healthcare Scientist | 1.31 | 0.66, 2.59 | 0.45 | 1.47 | 0.72, 2.97 | 0.29 |
| Pharmacist | 0.57 | 0.13, 2.51 | 0.46 | 0.53 | 0.12, 2.39 | 0.41 |
| Therapist <sup>a</sup> | 0.88 | 0.31, 2.5 | 0.82 | 1.09 | 0.38, 3.16 | 0.88 |
| Student | 1.77 | 0.67, 4.67 | 0.25 | 1.53 | 0.57, 4.10 | 0.40 |
| Estates/Porters | 1.23 | 0.25, 5.98 | 0.80 | 1.11 | 0.22, 5.61 | 0.90 |
| Other | 1.34 | 0.71, 2.54 | 0.37 | 1.32 | 0.68, 2.56 | 0.42 |
| **Medical group** |  |  |  |  |  |  |
| No medial conditions | Ref |  |  | Ref |  |  |
| Immunosuppression | 0.75 | 0.26, 2.14 | 0.59 | 0.83 | 0.28, 2.45 | 0.74 |
| Chronic respiratory conditions | 1.23 | 0.78, 1.94 | 0.36 | 1.26 | 0.79, 2.00 | 0.33 |
| Chronic non-respiratory conditions | 0.88 | 0.52, 1.49 | 0.64 | 0.78 | 0.45, 1.37 | 0.39 |

*Note: Not vaccinated defined as not receiving the COVID-19 Autumn 2023 vaccine; Vaccinated as defined as receiving the COVID-19 Autumn 2023 vaccine for at least 14 days*
<sup>a</sup> *Therapist includes: Physiotherapist, Occupational Therapist, and Speech and Language Therapists*

## References

1. Elliot AJ, Cross KW, Fleming DM. Acute respiratory infections and winter pressures on hospital admissions in England and Wales 1990-2005. J Public Health (Oxf). 2008 Mar;30(1):91–8. PubMed PMID: 18258786. Epub 20080207. eng.

2. Morbey RA, Charlett A, Lake I, Mapstone J, Pebody R, Sedgwick J, et al. Can syndromic surveillance help forecast winter hospital bed pressures in England? PLoS One. 2020;15(2):e0228804. PubMed PMID: 32040541. Pubmed Central PMCID: PMC7010388. Epub 20200210. eng.

3. Luu B, McCoy-Hass V, Kadiu T, Ngo V, Kadiu S, Lien J. Severe Acute Respiratory Syndrome Associated Infections. Physician Assist Clin. 2023 Jul;8(3):495–530. PubMed PMID: 37197227. Pubmed Central PMCID: PMC10015106. Epub 20230315. eng.

4. Richter VP, de-Paris F, Pires MR, Bock H. Epidemiology of respiratory viruses before and during the COVID-19 pandemic in a tertiary care hospital in Southern Brazil. Journal of Clinical Virology Plus. 2024 2024/11/01/;4(4):100190.

5. Chow EJ, Uyeki TM, Chu HY. The effects of the COVID-19 pandemic on community respiratory virus activity. Nature Reviews Microbiology. 2023 2023/03/01;21(3):195-210.

6. UK Health Security Agency. Influenza: the green book, chapter 19 www.gov.uk2013 [updated 10/11/202306/12/2024]. Available from: https://www.gov.uk/government/publications/influenza-the-green-book-chapter-19.

7. UK Health Security Agency. Respiratory syncytial virus: the green book, chapter 27a www.gov.uk2013 [updated 01/10/202406/12/2024]. Available from: https://www.gov.uk/government/publications/respiratory-syncytial-virus-the-green-book-chapter-27a.

8. UK Health Security Agency. COVID-19: the green book, chapter 14a www.gov.uk2020 [updated 16/09/202406/12/2024]. Available from: https://www.gov.uk/government/publications/covid-19-the-green-book-chapter-14a.

9. Imai C, Toizumi M, Hall L, Lambert S, Halton K, Merollini K. A systematic review and meta-analysis of the direct epidemiological and economic effects of seasonal influenza vaccination on healthcare workers. PLoS One. 2018;13(6):e0198685. PubMed PMID: 29879206. Pubmed Central PMCID: PMC5991711. Epub 20180607. eng.

10. Wallace S, Hall V, Charlett A, Kirwan PD, Cole M, Gillson N, et al. Impact of prior SARS-CoV-2 infection and COVID-19 vaccination on the subsequent incidence of COVID-19: a multicentre prospective cohort study among UK healthcare workers - the SIREN (Sarscov2 Immunity & REinfection EvaluatioN) study protocol. BMJ Open. 2022 Jun 28;12(6):e054336. PubMed PMID: 35768083. Pubmed Central PMCID: PMC9240450. Epub 20220628. eng.

11. Broad J, Sparkes D, Platt N, Howells A, Foulkes S, Khawam J, et al. Adapting COVID-19 research infrastructure to capture influenza and respiratory syncytial virus alongside SARS-CoV-2 in UK healthcare workers winter 2022/23 and beyond: protocol for a pragmatic sub-study [version 3; peer review: 1 approved, 1 approved with reservations]. NIHR Open Research. 2024;4(1).

12. Russell S, Munro K, Foulkes S, Broad J, Sparkes D, Atti A, et al. Adapting COVID-19 research infrastructure to capture influenza and RSV alongside SARS-CoV-2 in UK healthcare workers winter 2022/23: Evaluation of the SIREN Winter Pressures pilot study. medRxiv. 2024:2024.09.08.24313279.

13. Foulkes S. Adapting COVID-19 research infrastructure to capture influenza and respiratory syncytial virus alongside SARS-CoV-2 in UK healthcare workers winter 2022/23: Results of a pilot study in the SIREN cohort. medRxiv. 2024.

14. Tonkin-Hill G, Martincorena I, Amato R, Lawson ARJ, Gerstung M, Johnston I, et al. Patterns of within-host genetic diversity in SARS-CoV-2. eLife. 2021 2021/08/13;10:e66857.

15. Wang J, Jiang L, Xu Y, He W, Zhang C, Bi F, et al. Epidemiology of influenza virus reinfection in Guangxi, China: a retrospective analysis of a nine-year influenza surveillance data: Characteristics of influenza virus reinfection. International Journal of Infectious Diseases. 2022 2022/07/01/;120:135-41.

16. Hall CB, Walsh EE, Long CE, Schnabel KC. Immunity to and frequency of reinfection with respiratory syncytial virus. J Infect Dis. 1991 Apr;163(4):693–8. PubMed PMID: 2010624. eng.

17. Hall VJ, Foulkes S, Charlett A, Atti A, Monk EJM, Simmons R, et al. SARS-CoV-2 infection rates of antibody-positive compared with antibody-negative health-care workers in England: a large, multicentre, prospective cohort study (SIREN). The Lancet. 2021;397(10283):1459–69.

18. Atti A, Ferrari M, Castillo-Olivares J, Monk EJM, Gopal R, Patel M, et al. Serological profile of first SARS-CoV-2 reinfection cases detected within the SIREN study. Journal of Infection. 2022;84(2):248–88.

19. UK Health Security Agency. Surveillance of influenza and other seasonal respiratory viruses in the UK, winter 2023 to 2024 Gov.uk2024 [updated 06/09/202405/12/2024]. Available from: https://www.gov.uk/government/statistics/surveillance-of-influenza-and-other-seasonal-respiratory-viruses-in-the-uk-winter-2023-to-2024/surveillance-of-influenza-and-other-seasonal-respiratory-viruses-in-the-uk-winter-2023-to-2024.

20. UK Health Security Agency. National flu and COVID-19 surveillance reports: 2023 to 2024 season www.gov.uk2023 [updated 05/07/202406/12/2024]. Available from: https://www.gov.uk/government/statistics/national-flu-and-covid-19-surveillance-reports-2023-to-2024-season.

21. Shang W, Kang L, Cao G, Wang Y, Gao P, Liu J, et al. Percentage of Asymptomatic Infections among SARS-CoV-2 Omicron Variant-Positive Individuals: A Systematic Review and Meta-Analysis. Vaccines (Basel). 2022 Jun 30;10(7). PubMed PMID: 35891214. Pubmed Central PMCID: PMC9321237. Epub 20220630. eng.

22. Leung NH, Xu C, Ip DK, Cowling BJ. Review Article: The Fraction of Influenza Virus Infections That Are Asymptomatic: A Systematic Review and Meta-analysis. Epidemiology. 2015 Nov;26(6):862–72. PubMed PMID: 26133025. Pubmed Central PMCID: PMC4586318. eng.

23. Ip DK, Lau LL, Leung NH, Fang VJ, Chan KH, Chu DK, et al. Viral Shedding and Transmission Potential of Asymptomatic and Paucisymptomatic Influenza Virus Infections in the Community. Clin Infect Dis. 2017 Mar 15;64(6):736–42. PubMed PMID: 28011603. Pubmed Central PMCID: PMC5967351. eng.

24. Lau LL, Cowling BJ, Fang VJ, Chan KH, Lau EH, Lipsitch M, et al. Viral shedding and clinical illness in naturally acquired influenza virus infections. J Infect Dis. 2010 May 15;201(10):1509–16. PubMed PMID: 20377412. Pubmed Central PMCID: PMC3060408. eng.

25. Hayward AC, Fragaszy EB, Bermingham A, Wang L, Copas A, Edmunds WJ, et al. Comparative community burden and severity of seasonal and pandemic influenza: results of the Flu Watch cohort study. Lancet Respir Med. 2014 Jun;2(6):445–54. PubMed PMID: 24717637. Pubmed Central PMCID: PMC7164821. Epub 20140317. eng.

26. Moreira LP, Watanabe ASA, Camargo CN, Melchior TB, Granato C, Bellei N. Respiratory syncytial virus evaluation among asymptomatic and symptomatic subjects in a university hospital in Sao Paulo, Brazil, in the period of 2009-2013. Influenza Other Respir Viruses. 2018 May;12(3):326-30. PubMed PMID: 29078028. Pubmed Central PMCID: PMC5907818. Epub 20180204. eng.

27. Kirwan PD, Foulkes S, Munro K, Sparkes D, Singh J, Henry A, et al. Protection of vaccine boosters and prior infection against mild/asymptomatic and moderate COVID-19 infection in the UK SIREN healthcare worker cohort: October 2023 to March 2024. J Infect. 2024 Nov;89(5):106293. PubMed PMID: 39343245. Epub 20240927. eng.

28. Savulescu C, Prats-Uribe A, Brolin K, Uusküla A, Bergin C, Fleming C, et al. Effectiveness of the autumn 2023 COVID-19 vaccine dose in hospital-based healthcare workers: results of the VEBIS healthcare worker vaccine effectiveness cohort study, seven European countries, season 2023/24. Eurosurveillance. 2024;29(44):2400680.

29. Foulkes S, Munro K, Sparkes D, Broad J, Platt N, Howells A, et al. Adapting COVID-19 research infrastructure to capture influenza and respiratory syncytial virus alongside SARS-CoV-2 in UK healthcare workers winter 2022/23: Results of a pilot study in the SIREN cohort. medRxiv. 2024:2024.12.09.24318698.

